# A randomized clinical trial of a booster dose with low versus standard dose of AZD1222 in adult after 2 doses of inactivated vaccines

**DOI:** 10.1101/2022.02.15.22270974

**Authors:** Sira Nanthapisal, Thanyawee Puthanakit, Peera Jaru-Ampornpan, Rapisa Nantanee, Pimpayao Sodsai, Orawan Himananto, Jiratchaya Sophonphan, Pintip Suchartlikitwong, Narin Hiransuthikul, Pornpimon Angkasekwinai, Auchara Tangsathapornpong, Nattiya Hirankarn, the study team

## Abstract

**Background:** Immunogenicity of inactivated SARS-CoV-2 vaccine has waning antibody over time. With the emergence of the SARS-CoV-2 delta variant, which requires higher neutralizing antibody to prevent infection, a booster dose is needed.

**Objective:** To evaluate immunogenicity and reactogenicity of standard- versus low-dose ChAdOx1 nCoV-19 vaccine booster after CoronaVac in healthy adults.

**Methods:** A double-blinded, randomized, controlled trial of adult, aged 18-59 years, with completion of 2-dose CoronaVac at 21-28 days apart for more than 2 months was conducted. Participants were randomized to receive AZD1222 (Oxford/AstraZeneca) intramuscularly; standard dose (SD, 5×10^10^ viral particles) or low dose (LD, 2.5×10^10^ viral particles). Surrogate virus neutralization test (sVNT) against wild type and delta variant, and anti-spike-receptor-binding-domain IgG (anti-S-RBD IgG) were compared as geometric mean ratio (GMR) at day 14 and 90 between LD and SD arms.

**Results:** From July-August 2021, 422 adults with median age of 44 (IQR 36–51) years were enrolled. The median interval from CoronaVac to AZD1222 booster was 77 (IQR 64–95) days. At baseline, geometric means (GMs) of sVNT against delta variant and anti-S-RBD IgG were 18.1%inhibition (95%CI 16.4-20.0) and 111.5 (105.1-118.3) BAU/ml. GMs of sVNT against delta variant and anti-S-RBD IgG in SD were 95.6%inhibition (95%CI 94.3-97.0) and 1975.1 (1841.7-2118.2) BAU/ml at day 14, and 89.4%inhibition (86.4-92.4) and 938.6 (859.9-1024.4) BAU/ml at day 90, respectively. GMRs of sVNT against delta variant and anti-S-RBD IgG in LD compared to SD were 1.00 (95%CI 0.98-1.02) and 0.84 (0.76–0.93) at day 14, and 0.98 (0.94-1.03) and 0.89 (0.79-1.00) at day 90, respectively. LD recipients had significantly lower rate of fever (6.8%vs25.0%) and myalgia (51.9%vs70.7%) compared to SD.

**Conclusion:** Half-dose AZD1222 booster after 2-dose inactivated SARS-CoV-2 vaccination had non-inferior immunogenicity, yet lower systemic reactogenicity. Fractional low-dose AZD1222 booster should be considered especially in resource-constrained settings.

**Highlights:**

- Low dose AZD1222 could boost comparable immunity to standard dose in healthy adult who completed 2 doses of inactivated SARS-CoV-2 vaccines.
- Less reactogenicity occurred in low-dose AZD1222 booster than standard-dose recipients.

Thai Clinical Trials Registry (thaiclinicaltrials.org): TCTR20210722003

## 1. Introduction

The coronavirus disease 2019 (COVID-19) pandemic, caused by severe acute respiratory syndrome coronavirus 2 (SARS-CoV-2), has global impacts, with over 330 million cases worldwide, and over 5 million deaths [1]. In Thailand, as of January 2022, more than 2.3 million people with COVID-19 were reported with over 21,000 mortalities [1]. Moreover, the circulating SARS-CoV-2 has shifted from wild type WA1/2020 to several variants of concern. Since May 2021, the B.167.2 (delta variant) was the major cause of outbreak in many countries all over the world, including Thailand [2]. Variants of concern contain amino acid changes in the receptor binding domain (RBD) of spike protein which is the vaccine antigen. The neutralizing activity of serum samples from vaccinated persons against B.167.2 variant was reduced, compared with WA1/2020 variant [3]. The change in virus and waning of immunity after vaccination are driving forces for the necessity of a booster dose vaccine. World Health Organization stated that COVID-19 vaccine booster doses might be needed, considering on waning immunity, lower vaccine effectiveness against variants of concern, and global vaccine coverage [4].

Multiple vaccine platforms against SARS-CoV-2, including inactivated vaccines, and viral vector vaccines, have been rolled out in Thailand since March 2021. The inactivated SARS-CoV-2 vaccine (CoronaVac, Sinovac Life Sciences, Beijing, China) was shown to be effective in protecting against severe COVID-19 and COVID-19-related death, with two-dose efficacy of 65.9% against COVID-19 and 86.3% against COVID-19–related death [5]. However, the efficacy of this vaccine gradually decreased during the extended follow-up period, as shown by the increasing incidence of symptomatic SARS-CoV-2 infection in immunized individuals [6] and waning immunity [7]. Furthermore, CoronaVac was shown to induce lower neutralizing antibodies against variants of concern [8]. The ChAdOx1 nCoV-19 vaccine (AZD1222, Oxford/AstraZeneca) comprises a replication-deficient chimpanzee adenoviral vector ChAdOx1, containing the SARS-CoV-2 structural surface glycoprotein antigen (spike protein; nCoV-19) gene [9]. The report of randomized controlled trials of AZD1222 showed overall vaccine efficacy of 90% and 70.4%, from low-dose priming group and standard dose group, respectively [9]. The concept of fractional low dose was also shown in several studies. A quarter dose of mRNA-1273 could generate spike-specific memory CD4+ T cells in all participants and spike-specific CD8+ T cells in 88% of participants at 6 months after 2-dose completion, which were comparable in quantity and quality to COVID-19 cases [10]. As a booster dose, half dose and one-fifth dose of mRNA-1273 could boost neutralization titers against wild type and beta variant at 1 month after booster doses, given at 6 months after mRNA-1273 primary vaccination series [11].

Heterologous prime-boost vaccinations, the sequential administration of vaccines using different antigen delivery systems [12], have been reported as a good strategy to enhance cellular immune response against various viral pathogens including SARS-CoV-2 [13]. Studies on heterologous prime-boost vaccinations against SARS-CoV-2, using lipid nanoparticle-formulated mRNA vaccine BNT162b2 (BioNTech/Pfizer) as a booster dose in AZD1222-primed participants, showed significantly higher frequencies of spike-specific CD4+ and CD8+ T cells than participants who received two-dose AZD1222 [14]. In CoronaVac followed by AZD1222 vaccinees, the geometric mean of spike receptor binding domain (S-RBD) IgG titers were higher than 2-dose AZD1222 vaccinees, with the level of 1492 BAU/ml and 178 BAU/ml, respectively [15]. Moreover, a recent study from Thailand [16] showed that standard and half dose of BNT162b2 boosters after 2-dose inactivated vaccine series increased both antibody and cellular immune response against SARS-CoV-2.

The objective of this study was to determine antibody response, cellular response, and reactogenicity of standard versus low dose AZD1222 as a booster dose after completion of 2-dose CoronaVac in healthy adult.

## 2. Methods

### 2.1 Study design and participants

This study took place at two clinical research centers: Faculty of Medicine, Thammasat University, Pathum Thani, and Chulalongkorn University Health Center, Faculty of Medicine, Chulalongkorn University, Bangkok, Thailand. This is a double-blinded, randomized, controlled trial. Healthy adult, age 18 – 59 years old, who completed two doses of CoronaVac for more than 60 days, with interval of 21 – 28 days were included in this study. Participants who received any immunosuppressants or blood products within 3 months or any vaccines within 2 weeks before study enrollment were excluded. All participants provided written informed consent prior to study enrollment.

This study was registered in Thai Clinical Trials Registry (thaiclinicaltrials.org, TCTR20210722003). Institutional review board of Faculty of Medicine, Thammasat University (MTU-EC-PE1-182/64) and Faculty of Medicine, Chulalongkorn University (IRB No. 600/64) approved this study.

### 2.2 Study procedures

The participants were stratified into two age strata, age 18 – 45 years and 46 – 59 years. The randomization process was performed using a block of two or four in sealed envelope. The participants were vaccinated in participant-blinded fashion, with intramuscular AZD1222 lot number A1009 and A10061, manufactured by Siam Bioscience Co., Ltd., 0.25 ml (2.5 × 10^10^ viral particles, low dose [LD]) or 0.5 ml (5 × 10^10^ viral particles, standard dose [SD]). The vaccination was performed by unblinded nurses. Therefore, the participants and blinded study team were not aware of the randomization arms. During the first 7 days after vaccination, participants recorded the solicited local and systemic reactogenicity in the diary. The solicited local and systemic reactogenicity included pain at injection site, swelling, erythema, fever, feverish, headache, fatigue, myalgia, arthralgia (Chulalongkorn University study site), vomiting, and diarrhea. Blood samples were collected prior to booster doses administration. The follow-up visits were scheduled at day 14, and/or day 28, and day 90 to collect solicited and unsolicited reactogenicity data and perform blood collection. All participants’ samples were tested for spike receptor binding domain IgG (anti-S-RBD IgG) and surrogate virus neutralization test (sVNT) against wild type and B.167.2 (delta variant).

The cell-mediated immunity (CMI) sub study, to evaluate T and B cell responses, was performed among 80 random participants, prior to vaccination, day 28, and day 90, using enzyme-linked immunospot (ELISpot) assay.

### 2.3 Immunogenicity outcomes

#### 2.3.1 Quantitative spike receptor binding domain IgG (anti-S-RBD IgG) ELISA

The ELISA protocol was adapted from Amanat *et al*. (2020) [17]. Briefly, diluted serum samples were incubated in 96-well plates coated with purified recombinant Myc-His-tagged S-RBD, residues 319-541 from SARS-CoV-2 (Wuhan-Hu-1). Then, ELISA was performed. Anti-S-RBD IgG level was reported in binding-antibody units (BAU/mL) following conversion of OD450 values with the standard curve using known units of WHO international standard (NIBSC 20/136). We used anti-S-RBD IgG level at 506 BAU/ml, which is correlated with 80% vaccine efficacy reported by the Oxford COVID vaccine trial group [18], as a cut off.

#### 2.3.2 Surrogate virus neutralization test (sVNT)

A surrogate virus neutralization test was set up as previously described in Tan *et al*. (2020) [19]. Recombinant SRBD from the wild-type (Wuhan-Hu-1) and delta (B.1.617.2) strains were used. Serum samples (at 1:10 dilution) - SRBD mixture were incubated in 96-well plates coated with 0.1 µg/well recombinant human ACE2 ectodomain (GenScript). Then, ELISA was performed. The negative sample was pre-2019 human serum. The % inhibition was calculated as follows:

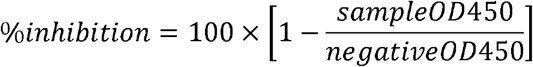

#### 2.3.3 Enzyme-linked immunospot (ELISpot) assay to evaluate T and B cell responses

**T cell responses** were assessed by ELISpot assay using a Human IFN-γ ELISpotPro™ kit (Mabtech, Stockholm, Sweden). Fresh peripheral blood mononuclear cells (PBMCs) (250,000 cells per well) were stimulated with overlapping peptide pools from 100 peptides of SARS-CoV-2 Spike (S) defined peptides and 101 peptides from the nucleoprotein (N), membrane protein (M) and open reading frame proteins (O) (Mabtech, Stockholm, Sweden) for 20 hours. Negative control and positive control, anti-CD3, were also included.

**B cell responses** were assessed by Human IgG SARS-CoV-2 RBD ELISpot PLUS (ALP) kit (Mabtech, Stockholm, Sweden). Briefly, fresh PBMCs (500,000 cells per well) were pre-stimulated with R848 and IL-2 for 72 hours to allow memory B cells to differentiate into antibody-secreting cells. Unstimulated well was also used as negative control. An RBD-WASP antigen was added into RBD-specific IgG detected well while MT78/145-biotinylated antibodies were added into total IgG detected well. Anti-WASP-ALP and streptavidin-ALP were added into RBD-specific IgG detected well and total IgG detected well, respectively.

The spots were counted using ImmunoSpot analyzer. Spot counts for negative control wells were subtracted from the test wells to generate normalized readings, these are presented as spot forming unit (SFU) per million PBMCs.

### 2.4 Reactogenicity

Solicited reactogenicity were assessed during the first 7-day period after booster vaccination, by self-recording diary. Grading of adverse events are checked according to the Guidance for Industry Toxicity Grading Scale for Healthy Adult and Adolescent Volunteers Enrolled in Preventive Vaccine Clinical Trials, 2007 [20]. Grading scale was grade 0 for no symptoms; grade 1 for mild symptom, which was not interfere with activities or vomiting 1 – 2 times/day or diarrhea 2 – 3 times/day; grade 2 for moderate symptom, which interfered with activities or needed medication for symptom relief, or vomiting more than 2 times/day or diarrhea 4 – 5 times/day; grade 3 for severe symptom, which incapacitated or diarrhea 6 or more times/day; grade 4 for potentially life-threatening symptom which required emergency room visit or hospitalization. Fever was graded as grade 1 (38.0 – 38.4°C), grade 2 (38.5 – 38.9°C), grade 3 (39 – 40°C), and grade 4 (more than 40°C). Feverish was defined as feeling of fever but body temperature less than 38.0°C. Unsolicited adverse events were also recorded by study team at all visits.

### 2.5 Statistical analysis

The sample size was calculated using a non-inferiority criterion for the geometric mean ratio (GMR) of sVNT against wild type and delta variant, and anti-S-RBD IgG, comparing LD to SD. Assuming 0.75 non-inferiority margin, 90% power, and one-sided statistical testing with 5% significance level, a minimum of 165 participants per group were required. Accounting for potentially missing data, the sample size was increased by 20%, yielding a total of 400 participants.

The demographic, laboratory data, and other continuous variables were described using median (interquartile range [IQR]), while the categorical variables were described using number with percentage. Wilcoxon rank sum test, chi-square or Fisher’s exact test were calculated to determine the differences in continuous and categorical variables between two groups, respectively. The primary endpoints, GMR of sVNT against wild type and delta variant, and anti-S-RBD IgG, comparing LD to SD at day 14 and 28 after booster, were compared in terms of non-inferiority. Non-inferiority was established if the lower bound of 95% confidence interval (CI) of GMR was greater than 0.67 [21]. The geometric means (GMs) were calculated as exponentiated means of logarithmic transformation of the assay results. Two-sided 95% CIs were antilogarithm of titers from difference of two-independent t-test. Anti-S-RBD IgG GM of at least 506 BAU/ml, the level corresponding with 80% vaccine efficacy against symptomatic infection [18], was used as the protective cut-off level. The reactogenicity rates were compared using chi-square test. All reported p-values are two-sided. P values <0.05 were considered to be statistically significant. Statistical analysis was performed using Stata version 15.1 (Stata Corp., College Station, Texas).

Correlation between the levels of anti-S-RBD IgG and receptor-blocking antibodies as measured by sVNT was analyzed for both wild type and delta strains by non-linear regression fit in GraphPad Prism 9 (GraphPad software, San Diego, CA). Data from the sub-population with anti-S-RBD IgG titer below 1200 BAU/mL were used in the correlation analysis.

## 3. Results

### 3.1 Baseline characteristics

From July to August 2021, 422 adults with median age of 44 (IQR 36 – 51) years participated. The trial profiles and demographic data are shown in Figure 1 and Table 1. The median interval from 2^nd^ dose of CoronaVac to AZD1222 booster dose was 77 days (IQR 64 – 95). Female was majority of the participants (50.7% in LD vs 53.6% in SD). Medians body mass index (BMI) of participants were 24.7 kg/m^2^ (IQR 21.9-27.7) in LD group and 24.8 kg/m^2^ (IQR 22 – 28.1) in SD group. Approximately one-third of participants had comorbidities e.g., hypertension, dyslipidemia, and diabetes mellitus. SD and LD groups had similar GMs of sVNT to delta variant and wild type, with 18.1% inhibition (95% CI 16.4 – 20.0) and 36.0% inhibition (95% CI 33.6 – 38.6), respectively. GMs of anti-S-RBD IgG were also comparable, with the level of 111.5 BAU/ml (95% CI 105.1 – 118.3).

**Table 1.**
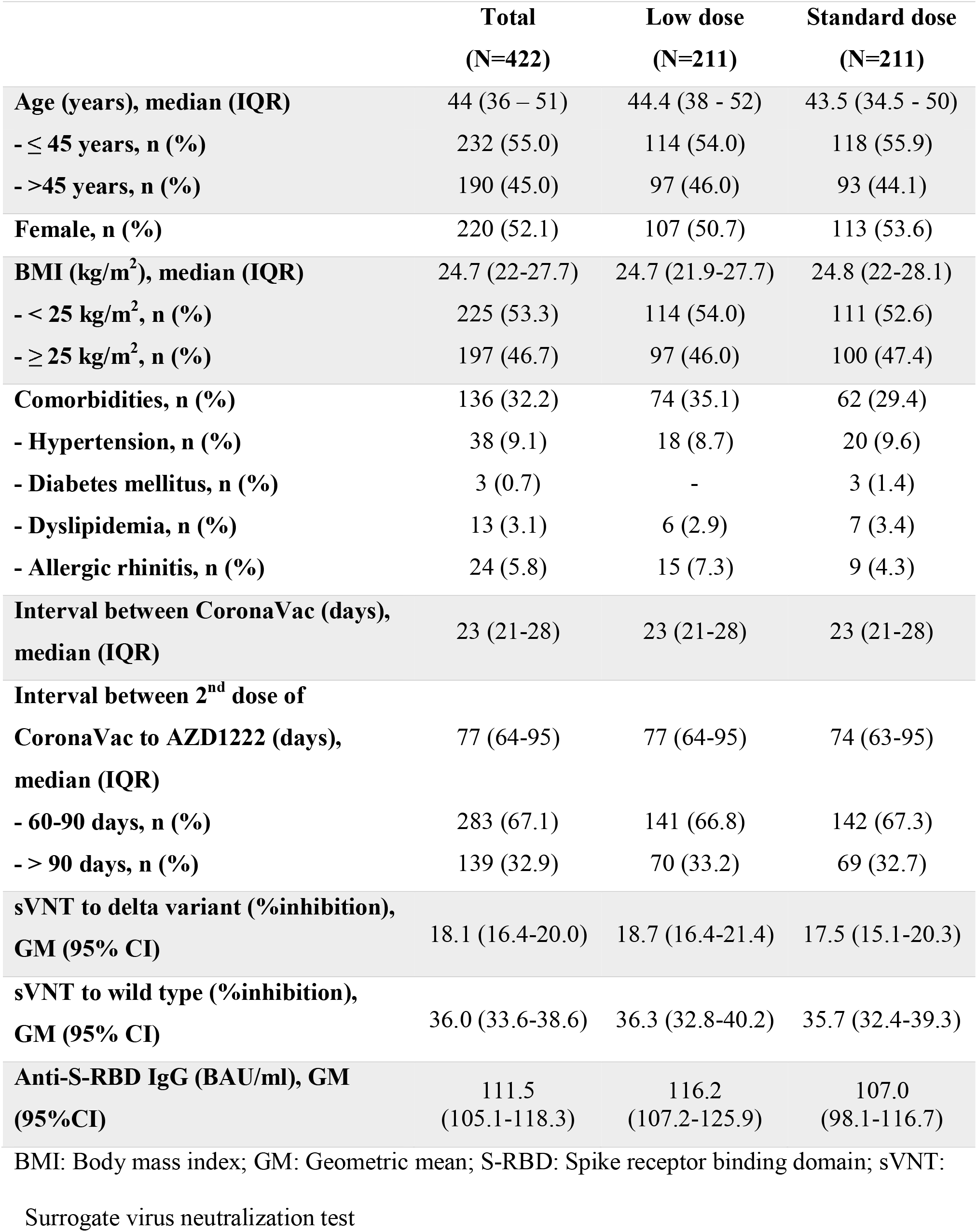
Baseline characteristics of study participants

|  | **Total**<br>**(N=422)** | **Low dose**<br>**(N=211)** | **Standard dose**<br>**(N=211)** |
| --- | --- | --- | --- |
| **Age (years), median (IQR)** | 44 (36 – 51) | 44.4 (38 - 52) | 43.5 (34.5 - 50) |
| **- ≤ 45 years, n (%)** | 232 (55.0) | 114 (54.0) | 118 (55.9) |
| **- >45 years, n (%)** | 190 (45.0) | 97 (46.0) | 93 (44.1) |
| **Female, n (%)** | 220 (52.1) | 107 (50.7) | 113 (53.6) |
| **BMI (kg/m<sup>2</sup>), median (IQR)** | 24.7 (22-27.7) | 24.7 (21.9-27.7) | 24.8 (22-28.1) |
| **- < 25 kg/m<sup>2</sup>, n (%)** | 225 (53.3) | 114 (54.0) | 111 (52.6) |
| **- ≥ 25 kg/m<sup>2</sup>, n (%)** | 197 (46.7) | 97 (46.0) | 100 (47.4) |
| **Comorbidities, n (%)** | 136 (32.2) | 74 (35.1) | 62 (29.4) |
| **- Hypertension, n (%)** | 38 (9.1) | 18 (8.7) | 20 (9.6) |
| **- Diabetes mellitus, n (%)** | 3 (0.7) | - | 3 (1.4) |
| **- Dyslipidemia, n (%)** | 13 (3.1) | 6 (2.9) | 7 (3.4) |
| **- Allergic rhinitis, n (%)** | 24 (5.8) | 15 (7.3) | 9 (4.3) |
| **Interval between CoronaVac (days),**<br>**median (IQR)** | 23 (21-28) | 23 (21-28) | 23 (21-28) |
| **Interval between 2<sup>nd</sup> dose of**<br>**CoronaVac to AZD1222 (days),**<br>**median (IQR)** | 77 (64-95) | 77 (64-95) | 74 (63-95) |
| **- 60-90 days, n (%)** | 283 (67.1) | 141 (66.8) | 142 (67.3) |
| **- > 90 days, n (%)** | 139 (32.9) | 70 (33.2) | 69 (32.7) |
| **sVNT to delta variant (%inhibition),**<br>**GM (95% CI)** | 18.1 (16.4-20.0) | 18.7 (16.4-21.4) | 17.5 (15.1-20.3) |
| **sVNT to wild type (%inhibition),**<br>**GM (95% CI)** | 36.0 (33.6-38.6) | 36.3 (32.8-40.2) | 35.7 (32.4-39.3) |
| **Anti-S-RBD IgG (BAU/ml), GM**<br>**(95%CI)** | 111.5<br>(105.1-118.3) | 116.2<br>(107.2-125.9) | 107.0<br>(98.1-116.7) |

BMI: Body mass index; GM: Geometric mean; S-RBD: Spike receptor binding domain; sVNT:
Surrogate virus neutralization test

**Figure 1.**
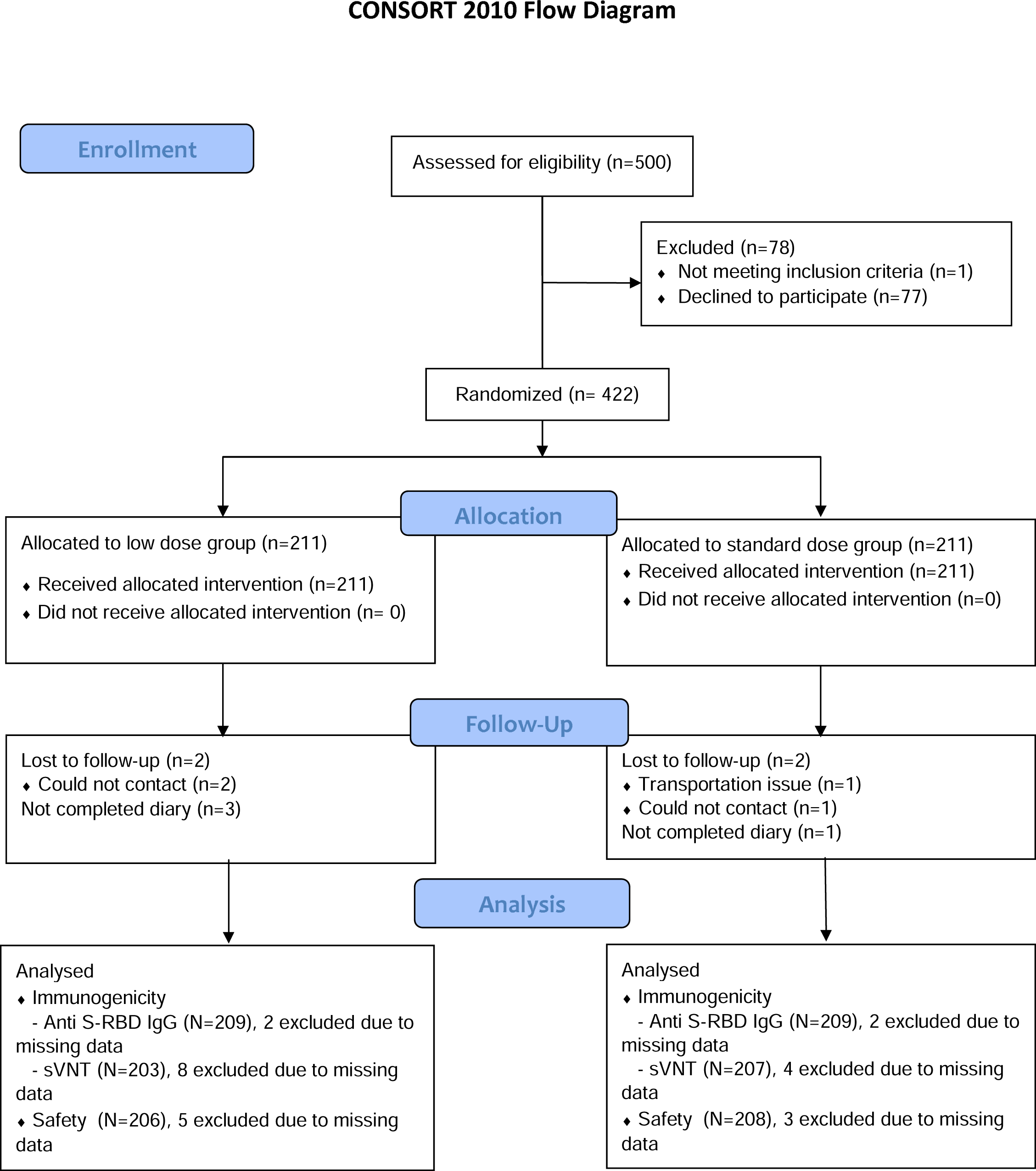
Participants disposition flow diagram

### 3.2 Reactogenicity

Proportions of participants who had local and systemic reactogenicities were shown in Figure 2 and Supplementary Table 1. LD recipients experienced significantly less systemic reactogenicity than SD recipients i.e., fever which defined as BT ≥ 38°C (LD, 6.8%; SD, 25%, p value <0.001), feverish (LD, 23.3%; SD, 38.9%, p-value <0.001), headache (LD, 54.9%; SD, 67.8%, p-value < 0.007), fatigue (LD, 63.1%; SD, 74.5%, p=0.01), and myalgia (LD, 51.9%; SD, 70.7%, p<0.001). Severe local and systemic reactogenicity or grade 3 events also occurred less frequently in LD, ranging from 0 – 5.3% in LD and 0 – 9.6% in SD, except vomiting. Grade 4 headache occurred in 1 SD participant, which improved by oral analgesics.

**Figure 2.**
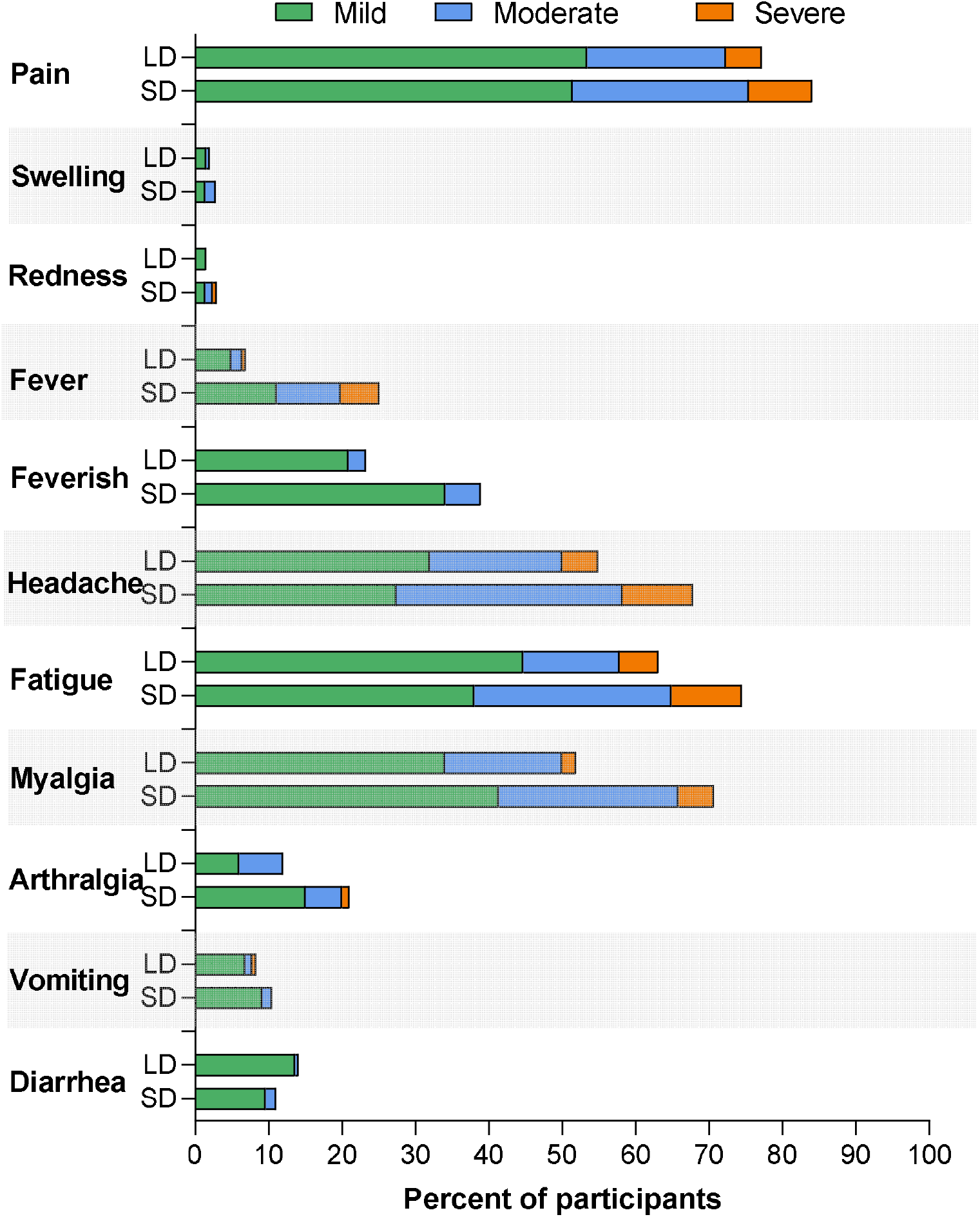
Local and systemic reactogenicities within 7 days after AZD1222 booster in adult after doses of inactivated vaccines, according to low dose and standard dose group. Note: LD= low dose AZD (2.5 × 10^10^ viral particles), SD= standard dose (5 × 10^10^ viral particles) The systemic reactions that are significant lower in the AZD1222 booster lower dose arm include: fever and feverish (p<0.001), headache (p<0.007), fatigue (p =0.01) and myalgia (p<0.001).

### 3.3 Immunogenicity

#### 3.3.1 SARS-CoV-2 neutralizing antibody by surrogate virus neutralization test (sVNT)

At 14 days post AZD1222 boosters, GMs of sVNT to delta variant were 95.6% inhibition (95% CI 94.6 – 96.7) in LD group and 95.6% inhibition (94.3 – 97.0) in SD group, as shown in Table 2 and Figure 3A. After 90 days, the GMs slightly decreased with the results of 87.9% inhibition (95% CI 85.4 – 90.5) in LD group and 89.4% inhibition (86.4 – 92.4) in SD group. GMR of sVNT against delta variant of LD to SD showed non-inferiority at both day 14 and day 90, with the value of 1.00 (95% CI 0.98 – 1.02) and 0.98 (0.94 – 1.03), respectively.

**Table 2.** Results of sVNT to delta variant, sVNT to wild type, and anti-S-RBD IgG at day 14 and day 90, and non-inferior comparison between low dose and standard dose AZD1222 booster in adult after 2 doses of inactivated vaccines.

| **Immunogenicity outcomes** | **Low dose**<br>**GM (95% CI)** | **Standard dose**<br>**GM (95% CI)** | **GMR**<br>**(95% CI)**<br>**LD to SD** |
| --- | --- | --- | --- |
| **sVNT to delta variant (%inhibition)** |  |  |  |
| **Day 14** | (N=203)<br>95.6 (94.6-96.7) | (N=207)<br>95.6 (94.3-97.0) | (N=410)<br>1.00 (0.98-1.02) |
| **Day 90** | (N=200)<br>87.9 (85.4-90.5) | (N=202)<br>89.4 (86.4-92.4) | (N=402)<br>0.98 (0.94-1.03) |
| **sVNT to wild type (%inhibition)** |  |  |  |
| **Day 14** | (N=203)<br>96.5 (95.8-97.1) | (N=207)<br>95.6 (93.3-97.8) | (N=410)<br>1.01 (0.99-1.03) |
| **Day 90** | (N=200)<br>92.1 (90.0-94.3) | (N=202)<br>93.1 (91.0-95.2) | (N=402)<br>0.99 (0.96-1.02) |
| **Anti-S-RBD IgG (BAU/ml)** |  |  |  |
| **Day 14** | (N=209)<br>1663.5 (1552.7-1782.2) | (N=209)<br>1975.1 (1841.7-2118.2) | (N=418)<br>0.84 (0.76-0.93) |
| **Day 90** | (N=206)<br>832.6 (767.9-902.8) | (N=203)<br>938.6 (859.9-1024.4) | (N=409)<br>0.89 (0.79-1.00) |

GM: Geometric mean; GMR: Geometric mean ratio; LD: Low dose; SD: Standard dose; S-RBD:
Spike receptor binding domain; sVNT: Surrogate virus neutralization test

**Figure 3A.**
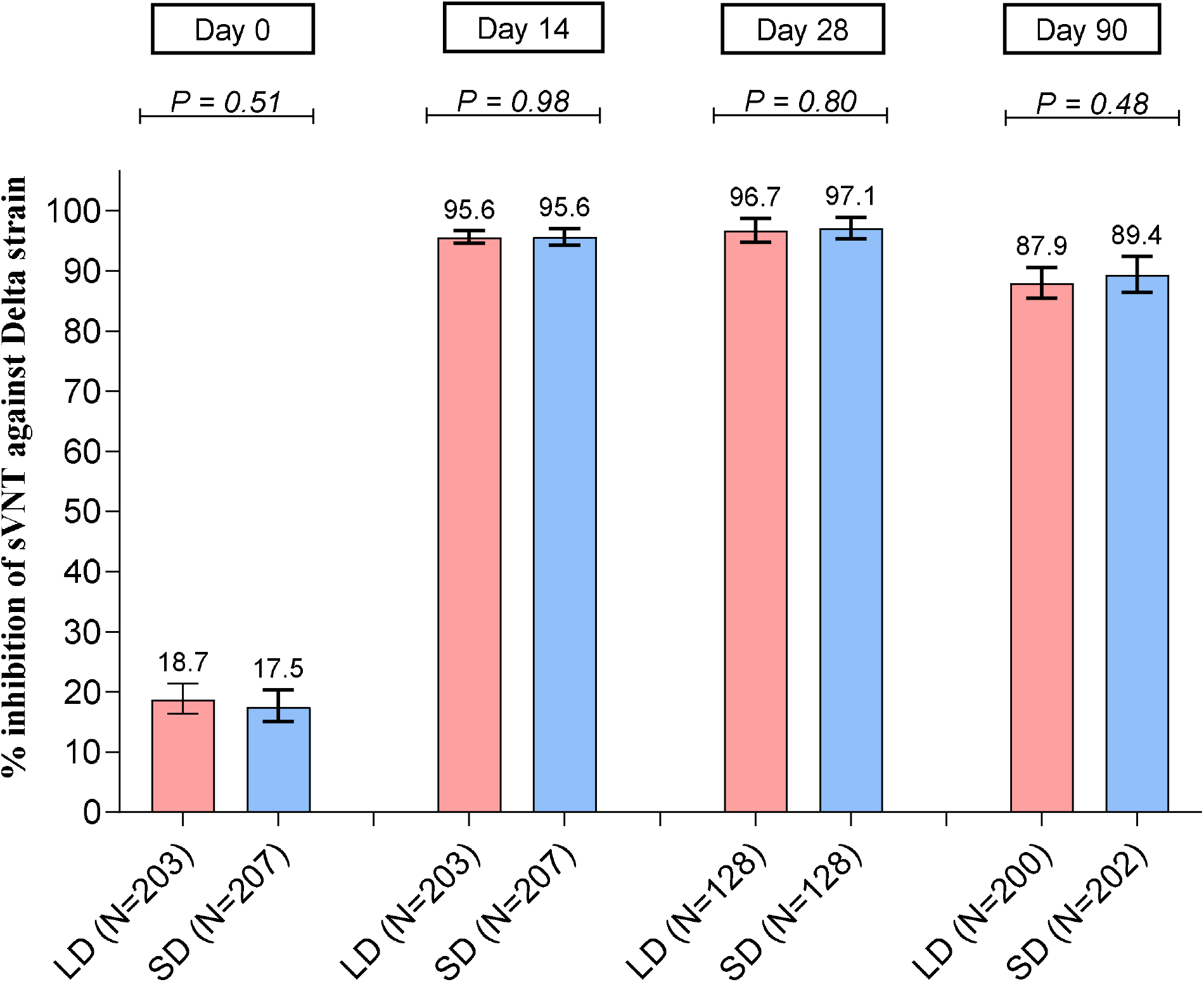
Geometric means (95% CI) of sVNT to delta variant of SARS-CoV-2, at day 0 (before AZD1222 booster doses), day 14, day 28, and day 90 after booster doses in adult after 2 doses of inactivated vaccines. P-value was evaluated by two-independent t-test. LD: Low dose; SD: Standard dose; sVNT: Surrogate virus neutralization test

**Figure 3B.**
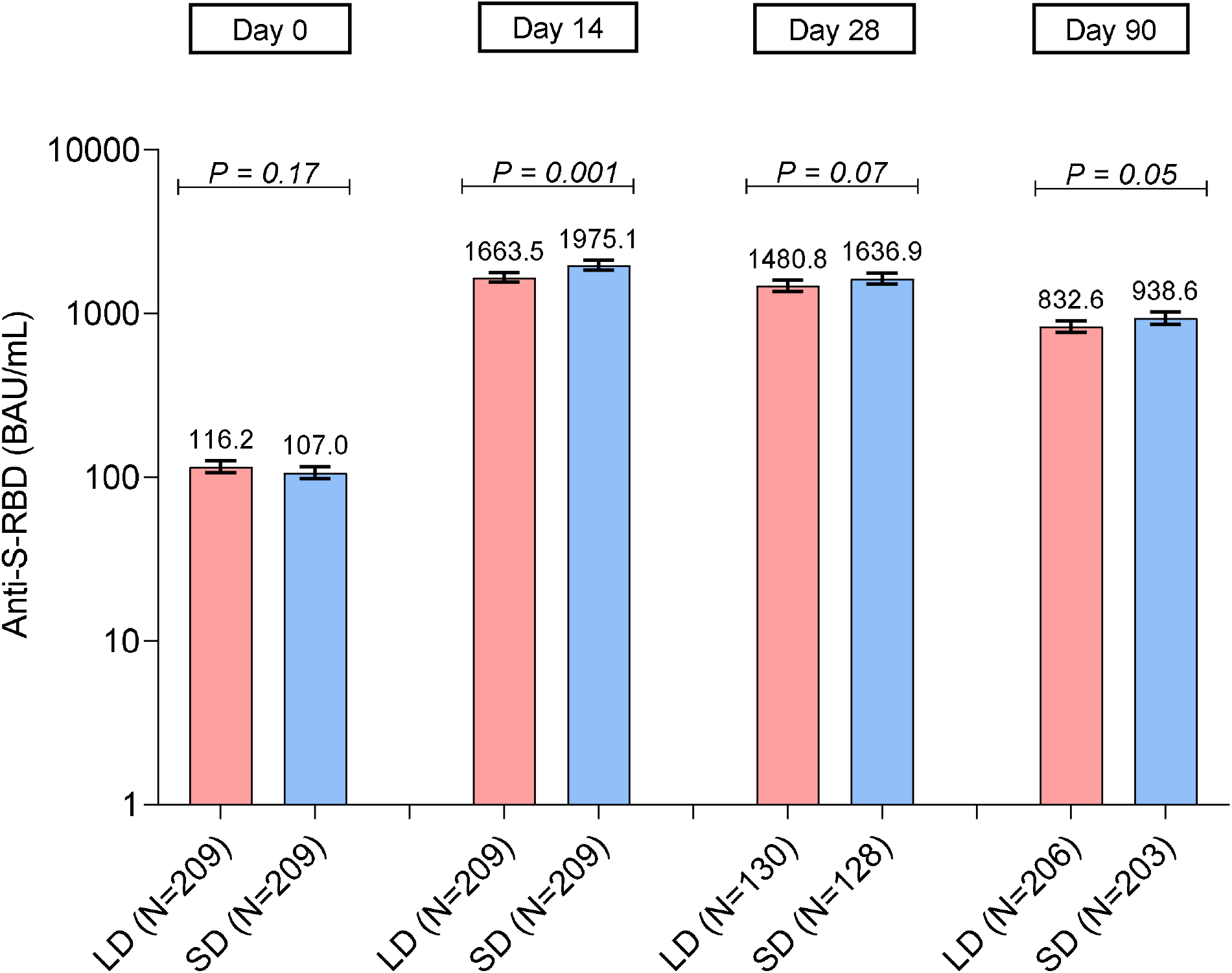
Geometric means (95% CI) of anti-S-RBD IgG (BAU/ml), at day 0 (before AZD1222 booster doses), day 14, day 28, and day 90 after booster doses in adult after 2 doses of inactivated vaccines. P-value was evaluated by two-independent t-test. LD: Low dose; SD: Standard dose; S-RBD: Spike receptor binding domain

GMs of sVNT to wild type were slightly higher than to delta variant, with 96.5% inhibition (95% CI 95.8 – 97.1) in LD group and 95.6% inhibition (93.3 – 97.8) in SD group at 14 days after booster, as shown in Table 2. At day 90, the GMs were 92.1% inhibition (95% CI 90.0 – 94.3) in LD group and 93.1% inhibition (91.0 – 95.2) in SD group, showing similar trend as sVNT against delta variants. Non-inferior GMRs were also concluded at both day 14 and day 90.

#### 3.3.2 SARS-CoV-2 binding antibody by anti-S-RBD IgG

At day 14 post booster doses, anti-S-RBD IgG GMs were increased to 1663.5 BAU/ml (95% CI 1552.7 – 1782.2) in LD and 1975.1 BAU/ml (1841.7 – 2118.2) in SD, as shown in Table 2 and Figure 3B. After 90 days, anti-S-RBD IgG GMs of both groups declined, with 832.6 BAU/ml (95% CI 767.9 – 902.8) in LD and 938.6 BAU/ml (859.9 – 1024.4) in SD, however, the levels were still above 506 BAU/ml, the level correlating with 80% vaccine efficacy against symptomatic infection [18]. GMR of anti-S-RBD IgG demonstrated non-inferiority of LD to SD at both day 14 and day 90. Subgroup analyses of participants’ characteristics on anti-S-RBD IgG at day 90; the results denoted the statistically significant higher among SD group compared with LD group among participants in age > 45 years (SD: 1144BAU/ml (95%CI 683-1608) versus LD: 851BAU/ml (95%CI 556-1324), p 0.01), and BMI ≥ 25 kg/m^2^ (SD: 1245 BAU/ml (95% CI 654 – 1628) vs LD: 875 (95% CI 626 – 1309), p 0.03), respectively.

### 3.4 Correlation between SARS CoV2 binding antibody and neutralization titer

The levels of antibodies capable of inhibiting binding between hACE2 receptor and S-RBD from both Wuhan and Delta strains were well-correlated with the levels of anti-RBD IgG (R^2^ = 0.80 for wild type and R^2^ = 0.84 for delta strain). Correlation analyses between anti-S-RBD IgG and sVNT from participants with anti-S-RBD less than 1200 BAU/ml at any time points were shown in Figure 4. Only 8.1% of participants with anti-RBD IgG levels higher than 1200 BAU/mL achieved lower than 95% inhibition levels in both sVNT assays. From non-linear fits of data, the levels of anti-S-RBD IgG that gave 68% and 80% inhibition against the wild type strain were approximately 298 (95% CI 291 – 305) and 472 (95% CI 452 – 494) BAU/mL, respectively. For the delta strain, these approximated anti-RBD IgG levels were slightly higher at 498 (95% CI 480 – 518) and 742 (95% CI 704 – 786) BAU/mL, respectively. The anti-S-RBD IgG level of 506 BAU/ml, the level correlating with 80% vaccine efficacy against symptomatic infection during the Alpha (B.1.1.7) variant outbreak in United Kingdom [18], is correlated with 81.8% inhibition of sVNT against wild type and 71% inhibition of sVNT against delta variant.

**Figure 4.**
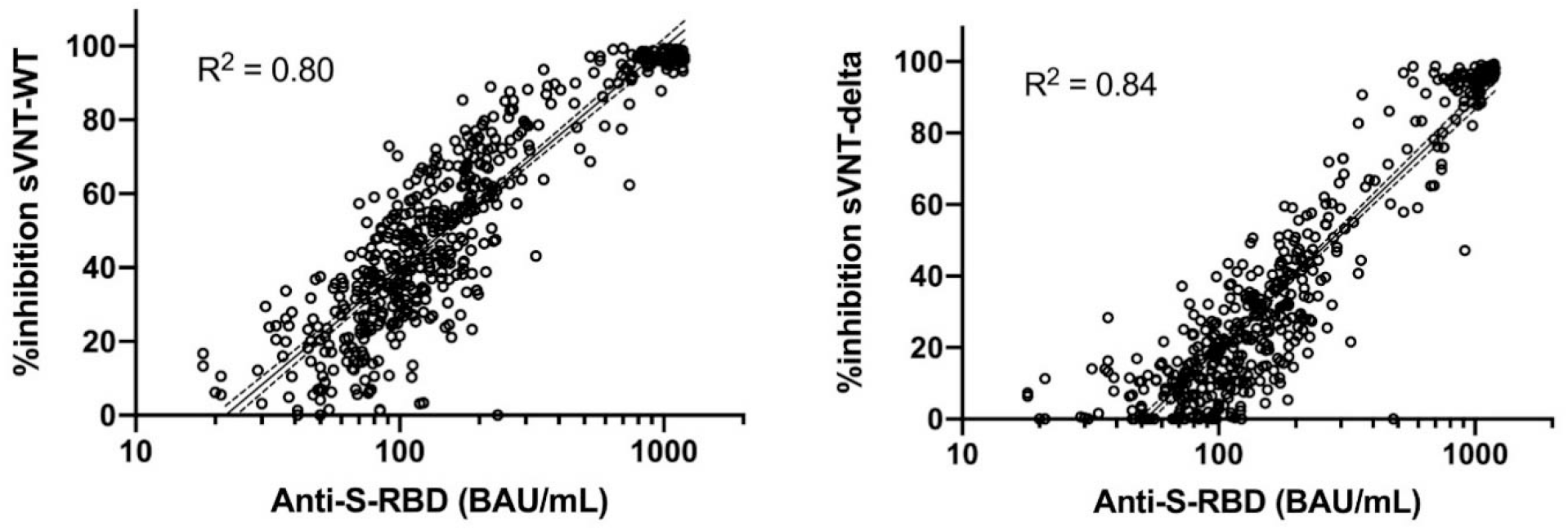
Correlation between anti-S-RBD IgG and sVNT to wild type (left) and delta variant (right) of SARS-CoV-2 at prior and post AZD1222 booster vaccination in adult after 2 doses of inactivated vaccines if anti-S-RBD IgG < 1200 BAU/ml. Dash lines represent 95% CI of fits of data. S-RBD: Spike receptor binding domain, sVNT: Surrogate virus neutralization test, WT: Wild type

### 3.5 Cell-mediated immune response by ELISpot assay

The median frequencies of T cell response to SNMO peptides and RBD-specific B cells with SARS-CoV-2 at baseline, and day 28 and day90 after booster dose are shown in Figure 5. At baseline prior to AZD1222 booster, T cell response to SNMO peptides were 60 (IQR 32 – 100) cells/10^6^ PBMCs in LD and 50 (20 -84) cells/10^6^ PBMCs in SD. At day 28 after receipt of booster, the median frequencies of SNMO-specific T cell response increased to 104 (IQR 56 - 196) and 80 (40 - 108) cells/10^6^ PBMCs in LD and SD group, respectively. The median frequencies of RBD-specific B cells were very low at baseline; 1 (IQR 0 - 16) cells/10^6^ PBMCs in LD and 0 (0 - 6.5) cells/10^6^ PBMCs in SD at day 0 and raised to 18 (8 - 42) and 26 (4 - 54) cells/10^6^ PBMCs at day 28 in LD and SD group, respectively. Both T and B cell responses were comparable between LD and SD group.

**Figure 5.**
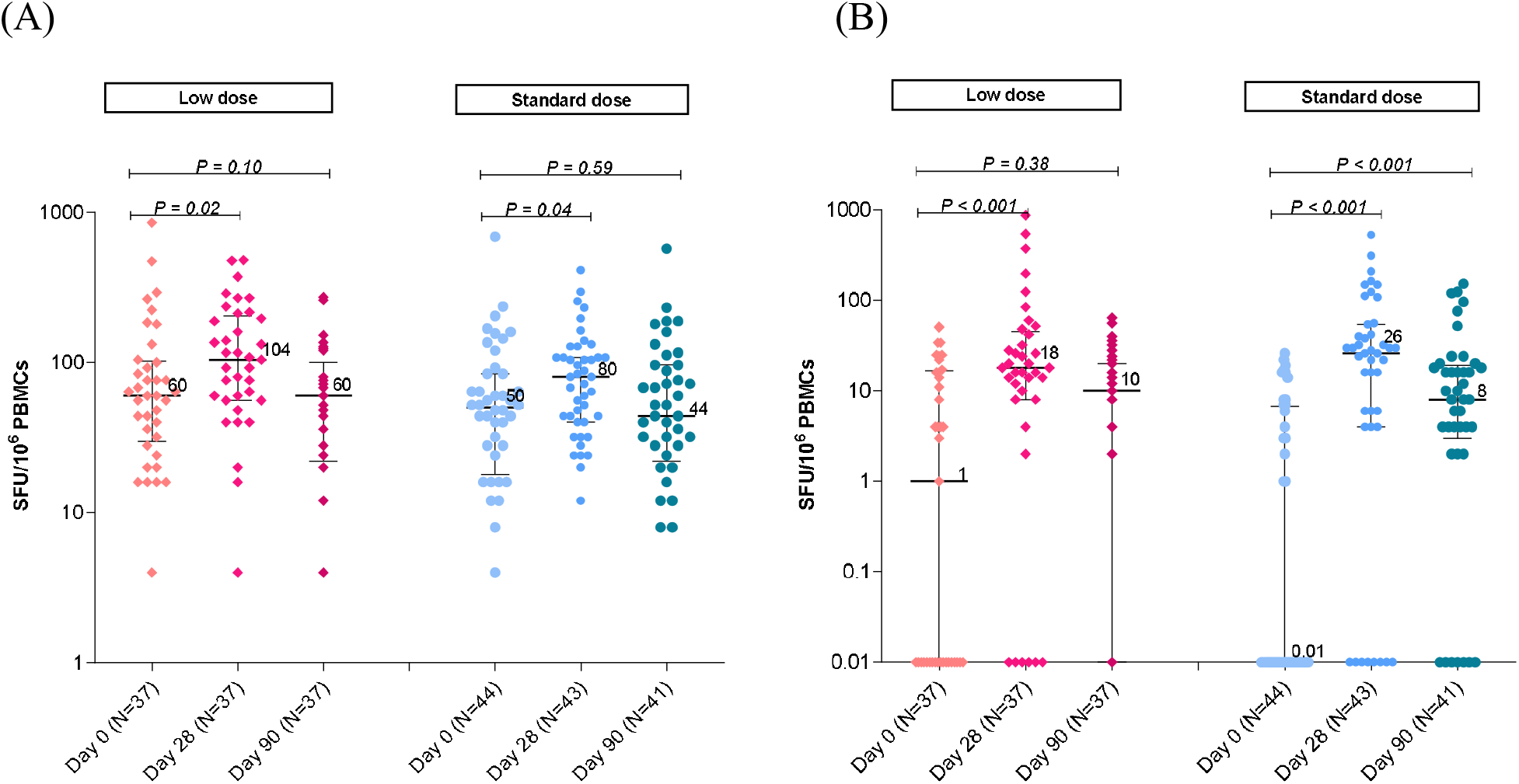
ELISpot assay at day 28 post AZD1222 booster in adult after 2 doses of inactivated vaccines, in low dose and standard dose group: (A) SNMO-specific T cell response, (B) RBD-specific B cell response. RBD: Receptor binding domain; SFU: spot forming unit; SNMO: Spike (S) nucleoprotein (N), membrane protein (M), and open reading frame proteins (O) of SARS-CoV-2

## 4. Discussion

The results showed that AZD1222 booster doses were able to boost immune responses after 14 days to > 95% neutralizing inhibition against SARS-CoV-2 strains in previous CoronaVac vaccinees. Low dose AZD1222 via intramuscular route had non-inferior immunogenicity response to full dose of vaccine measured by anti-S-RBD IgG and surrogate neutralizing antibodies, to both wild type and delta variant strains. Systemic reactogenicities especially fever, headache, fatigue, and myalgia, developed in fewer LD participants than those in SD group.

Solicited local and systemic adverse reactions after AZD1222 were more common after the first doses [22]. Our study found higher rates of fever, feverish, headache, fatigue, and myalgia in SD recipients. In participants receiving LD, headache, fatigue, myalgia, and pain at injection site frequently developed than the previous report [22], yet lower than those receiving SD. The less adverse reactions after LD AZD1222 may lead to more vaccine acceptability, especially during this COVID-19 pandemic, when additional booster vaccination might be needed.

The fractional low-dose concept of SARS-CoV-2 vaccine has been investigated. A quarter dose of mRNA-1273 was able to generate T cell responses, both spike-specific memory CD4+ T cells and spike-specific CD8+ T cells, in almost all of the participants at 6 months after 2 doses, which were comparable in quantity and quality to COVID-19 cases [10]. In the view of immunological memory, lower antigen level could activate quicker and stronger secondary immune response to previously encountered antigen [23]. Therefore, low-dose vaccination could be an effective booster.

Feng et al. [18] reported that the anti-S-RBD IgG level associated with 80% vaccine efficacy against symptomatic COVID-19 was 506 BAU/ml. For sVNT, 68% inhibition might correlate with vaccine efficacy [24]. Based on these indicative cut-off levels, low dose AZD1222 booster doses, in previous CoronaVac recipients, were effective. In phase 2/3 trials of AZD1222, the low dose/standard dose cohort showed higher vaccine efficacy at 90%, compared to 70.4% in standard dose group [9]. These findings also support that low dose AZD1222 is the compelling choice if booster doses are needed.

Heterologous prime-boost schedule of COVID-19 vaccine was reported in many studies. The Com-COV trial showed that, at 28 days post boost, the geometric mean concentration of SARS-CoV-2 anti-spike IgG in AZD1222 followed by BNT162b2 schedule was higher than 2-dose AZD1222 schedule [25]. Moreover, other study [14] and systematic review [26] also reported robust immunogenicity of this heterologous administration of AZD1222 followed by BNT162b2. Currently, CoronaVac has been widely used, with more than 600 million doses delivered in more than 40 countries worldwide [27], with the emergence of SARS-CoV-2 variants, the booster dose after completion of inactivated SARS-CoV-2 vaccine schedule should be considered. The use of fractional low dose of COVID-19 vaccine could increase the vaccine coverage, while the vaccine supply is limited with only 15 percent vaccination coverage in Africa [28].

The strengths of this study were the double-blinded, randomized, controlled study design, and multiple methods of immunologic assessments, both humoral and cellular immunity, to wild type and variants of concern. We reported anti-S-RBD IgG and sVNT, especially to delta variant which was the major cause of outbreak in many countries, including Thailand. The limitation included the interval from last vaccination was only 2-3 months after completion of 2-dose CoronaVac. Though in the real world, the booster might be given at longer interval from last CoronaVac, in which we expected to have at least similar or even higher immunogenicity. Secondly, this study included healthy adults, and have no data among high-risk groups, such as the elderly or the immunocompromised patients.

Thirdly, the follow up of the participants are ongoing to observe the kinetic of antibody decline and risk of breakthrough infection if any occurred. Lastly, this study did not perform sVNT for recently emerged variants e.g., Omicron, however, the comparable T cell responses between standard and low dose AZD1222 booster could likely prevent severe symptoms of COVID-19 caused by these variants.

## Data Availability

All data produced in the present study are available upon reasonable request to the authors.

## Abbreviations

BAU: Binding-antibody unit
BMI: Body mass index
CMI: Cell-mediated immunity
ELISpot: Enzyme-linked immunospot
GM: Geometric mean
GMR: Geometric mean ratio
LD: Low dose
PBMC: Peripheral blood mononuclear cell
SFU: Spot forming unit
S-RBD: Spike receptor binding domain
SD: Standard dose
sVNT: Surrogate virus neutralization test

## Acknowledgements

Thank you for all study teams for their assistance with the study.

Faculty of Medicine, Thammasat University

Chulalongkorn University Health Service Center

Santhiti Dalan, MD, Supakanya Bintapanya, Kajeepat Rattanasrisompop, Pinyarat

Kitidumrongsook, Ratchadaporn Bamrungpipattanaporn, Nomjit Jirapattawong

Department of Microbiology, Faculty of Medicine, Chulalongkorn University

Pokrath Hansasuta, MD, Tysdi Laohasereekul, MD, Punyot Tovichayathamrong, MD,

Kasama Manothummetha, MD

Center of Excellence in Immunology and Immune-mediated Diseases, Department of Microbiology, Faculty of Medicine, Chulalongkorn University

Vichaya Ruenjaiman, PhD, Supapit Horpratum, Jullada Thawilwang

Center of Excellence in Pediatric Infectious Diseases and Vaccines, Department of Pediatrics, Faculty of Medicine, Chulalongkorn University

Puneyavee Aikphaibul, MD, Wipaporn Natalie Songtaweesin, MD, Tuangtip

Theerawit, Jitthiwa Athipunjapong, Thutsanum Meepuksom, Angsumalin Sutjarit,

Juthamanee Moonwong, Rachaneekorn Nadsasarn, Thidarat Jupimai, Pornpavee

Nuncharoen, Yanisar Chanpoom, Phattharapa Khamkhen

The HIV Netherlands Australia Thailand Research Collaboration (HIV-NAT), The Thai Red Cross AIDS Research Centre

Suwat Wongmueng, Palida Pingthaisong

National Center for Genetic Engineering and Biotechnology (BIOTEC)

Anan Jongkaewwattana, PhD, Kirana Yoohat, Channarong Seepiban, Jaraspim

Narkpuk, Thorntun Deangphare

---

**Supplementary Table 1.**
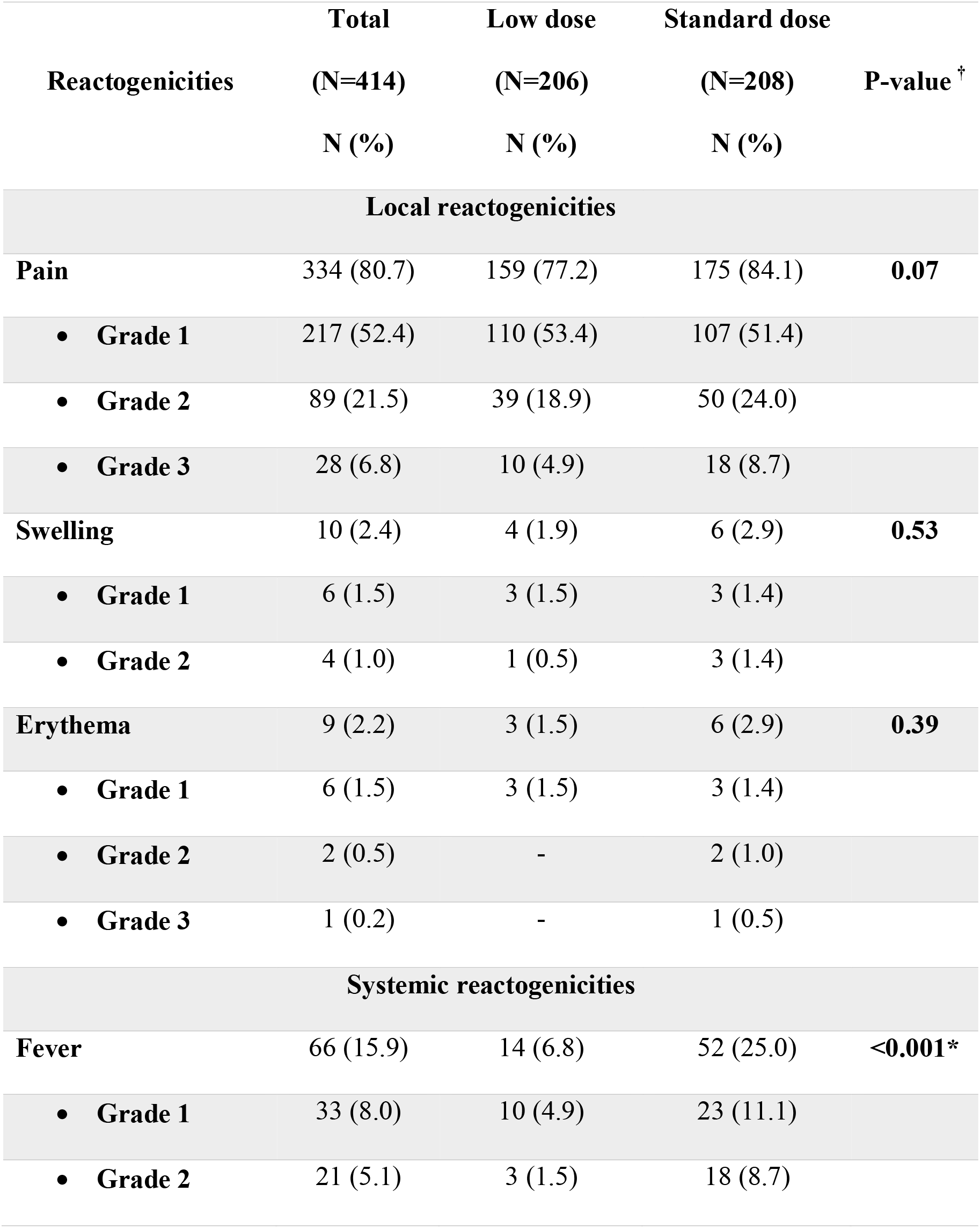
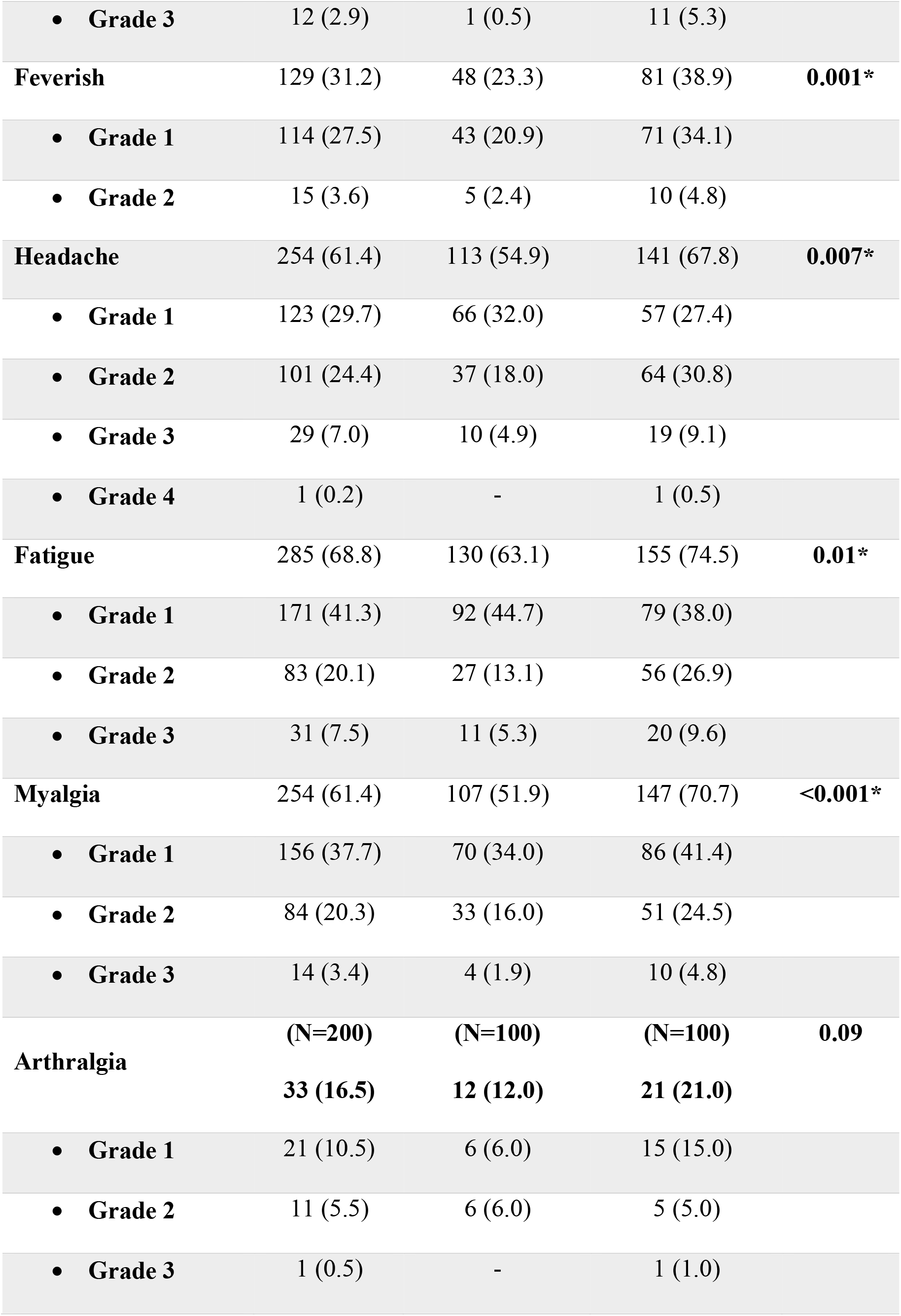
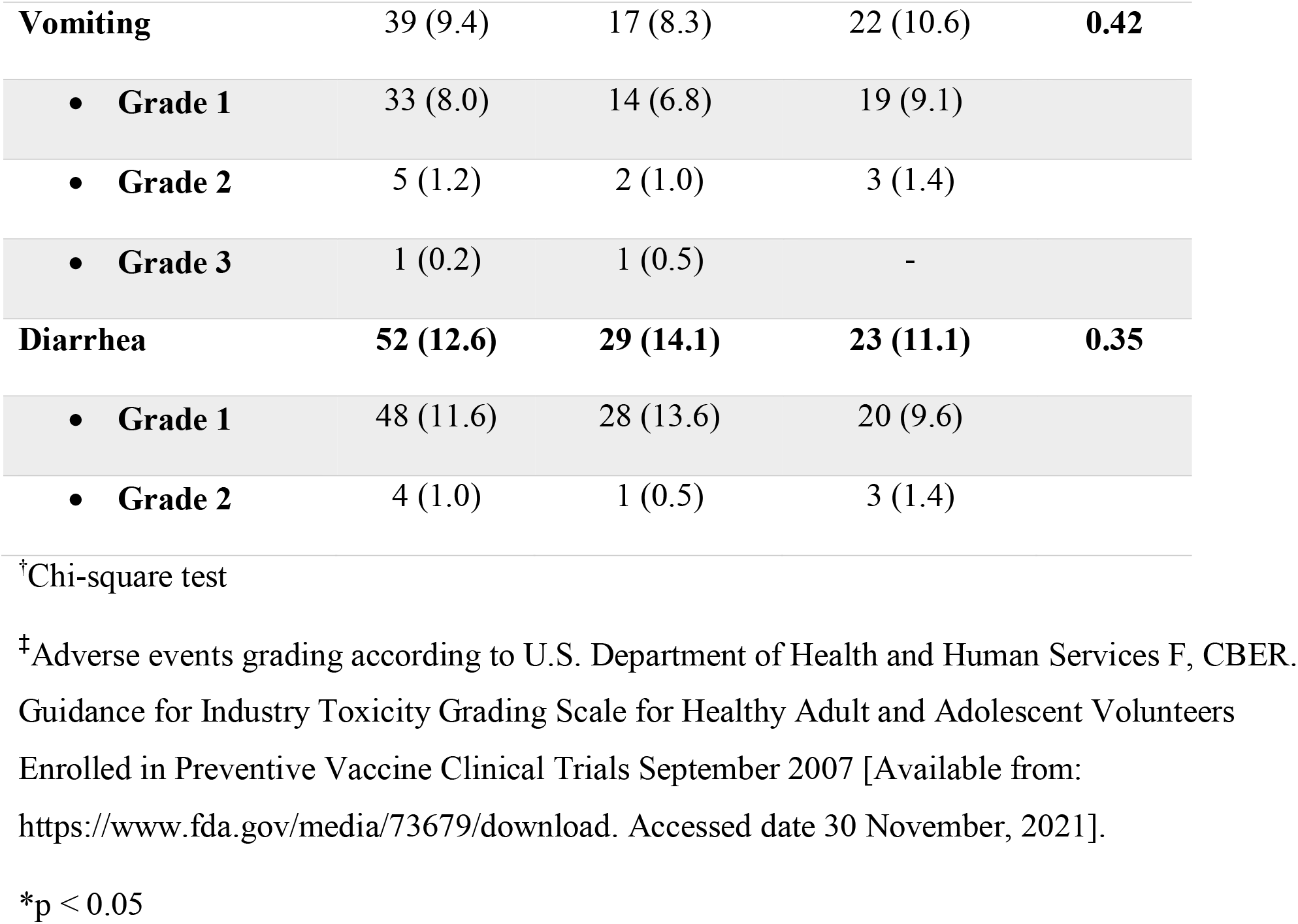
Local and systemic reactogenicities^‡^ within 7 days after AZD1222 booster in adult after 2 doses of inactivated vaccines, according to low dose and standard dose group.

